# Assessment of Commercial SARS-CoV-2 Antibody Assays, Jamaica

**DOI:** 10.1101/2020.09.27.20202655

**Authors:** Tiffany R. Butterfield, Alrica Bruce-Mowatt, Yakima Z. R. Phillips, Nicole Brown, Keisha Francis, Jabari Brown, Devon Taylor, Carl A. Bruce, Donovan McGrowder, Gilian Wharfe, Simone L. Sandiford, Tamara K. Thompson, Joshua J. Anzinger

## Abstract

The performance of the Roche Elecsys^®^ Anti-SARS-CoV-2, Abbott Architect SARS-CoV-2 IgG, Euroimmun SARS-CoV-2 IgA, Euroimmun SARS-CoV-2 IgG ELISA, and Trillium IgG/IgM rapid assays was evaluated in Jamaica, the largest country of the English-speaking Caribbean. Diagnostic sensitivities of the assays were assessed by testing serum samples from SARS-CoV-2 PCR-confirmed persons. Serum samples collected ≥14 days after onset of symptoms, or ≥14 days after an initial SARS-CoV-2 RT-PCR positive test for asymptomatics, showed diagnostic sensitivities ranging from 67.9-75.0% when including all possible disease severities and increased to 90.0-95.0% when examining those with moderate to critical disease. Grouping moderate to critical disease showed a significant association with a SARS-CoV-2 antibody positive result for all assays. Diagnostic specificity, assessed by testing serum samples collected during 2018-2019 from healthy persons and from persons with antibodies to a wide range of viral infections, ranged from 96.7-100.0%. For all assays examined, SARS-CoV-2 real-time PCR cycle threshold (Ct) values of the initial nasopharyngeal swab sample testing positive were significantly different for samples testing antibody positive versus negative. These data from a predominantly African descent Caribbean population shows comparable diagnostic sensitivities and specificities for all testing platforms assessed and limited utility of these tests for persons with asymptomatic and mild infections.

## 1. Introduction

The SARS-CoV-2 pandemic has resulted in an unprecedented need for reliable commercial laboratory diagnostics. While SARS-CoV-2 antibody assays have recently become commercially available, performance data have mainly assessed high-income country populations. Assessment of SARS-CoV-2 antibody assay performance in populations of mostly African descent is lacking. To our knowledge, there has been no published performance assessment of SARS-CoV-2 antibody assays with a predominantly black population. In this study in Jamaica, serum samples were used to assess the diagnostic sensitivity and specificity of five commercial SARS-CoV-2 antibody assays: Roche Elecsys^®^ Anti-SARS-CoV-2, Abbott Architect SARS-CoV-2 IgG, Euroimmun SARS-CoV-2 IgA and IgG ELISAs, and Trillium IgG/IgM rapid diagnostic test.

## 2. Materials and methods

For diagnostic sensitivity analysis, 42 blood samples collected in tubes without coagulant were obtained from 37 consenting persons testing SARS-CoV-2 real-time PCR positive. Disease severity was classified according to WHO criteria. Real-time PCR of nasopharyngeal swabs samples was performed at the Jamaica National Influenza Centre according to the Corman *et al* method [1]. Samples were collected 6-103 days after disease onset for symptomatic persons and 20-69 days after a positive real-time PCR test for asymptomatic persons.

For the assessment of diagnostic specificity, archival 2018-2019 serum samples from the University of the West Indies Virology Laboratory were identified. All samples were stored at −20°C. The specificity panel included serum samples testing positive for Zika virus IgM, chikungunya virus IgM, dengue virus IgM, CMV IgM, CMV IgG, EBV IgM, parvovirus B19 IgM, anti-HTLV-I/II, HIV Ag/Ab, HBsAg, anti-HCV, convalescent samples from patients seroconverting for influenza A or B virus antibodies, convalescent samples from patients with respiratory disease but without seroconversion for influenza A and B virus antibodies, healthy persons requesting vaccination status, and women seeking routine antenatal care (Supplementary Table).

The detection of antibodies was conducted with an Architect *i*2000SR for the Architect SARS-CoV-2 IgG assay, a cobas^®^ 6000 analyzer for the Elecsys^®^ Anti-SARS-CoV-2 assay, a Thermo Scientific Multiskan FC Microplate Photometer for the Euroimmun SARS-CoV-2 IgA and IgG ELISAs and lateral flow assay rapid test for the Trillium IgG/IgM assay. Each manufacturer’s instructions were used for cutoff index values for the Elecsys^®^, Architect, and Euroimmun platforms, and the appearance of a line of any intensity for the control and/or IgG and IgM for the Trillium IgG/IgM rapid test. Index values ≥1.0 are considered positive and index values <1.0 are considered negative for the Elecsys^®^ Anti-SARS-CoV-2 assay. For the Architect SARS-CoV-2 IgG assay index values ≥1.4 are considered positive and index values <1.4 are considered negative. For both Euroimmun SARS-CoV-2 ELISAs, index values ≥1.1 are considered positive, ≥0.8-<1.1 are considered borderline, and <0.8 are considered negative. Borderline index values were considered negative.

Data were analyzed using IBM SPSS Statistics^®^ for Windows version 20. Categorical variables were reported using proportions and continuous variables reported as mean with standard deviation. Comparison of means was by Welch’s t-test while associations between categorical variables were assessed using the χ^2^ test.

This study was approved by the UWI Mona Campus Research Ethics Committee (ECP 244 19/20).

## 3. Results

Diagnostic specificity of each assay was examined with a panel of archived serum samples predating the introduction of SARS-CoV-2 into Jamaica. The specificity was 100.0% (104/104) for Elecsys^®^ Anti-SARS-CoV-2, 98.2% (109/111) for Architect SARS-CoV-2 IgG, 97.5% (119/122) for Euroimmun SARS-CoV-2 IgA, 100.0% (122/122) for Euroimmun SARS-CoV-2 IgG, 96.7% (87/90) for Trillium IgM, and 98.9% (89/90) for Trillium IgG (Supplementary Table).

Diagnostic sensitivities of the assays ranged from 42.9-71.4% for samples collected 6-9 days after onset of symptoms, 85.7-100.0% for samples collected 10-13 days after onset of symptoms, and 90.0-95.0% for samples collected ≥14 days after onset of symptoms (Fig. 1). As samples from asymptomatic and mildly affected persons were only available for collections ≥14 days after onset of symptoms or after an initial SARS-CoV-2 PCR test, we examined sensitivities for all disease severities separately. For all assays, sensitivities were lower when asymptomatic and mild infections were included, ranging from 67.9-75.0% (Fig. 1). When moderate, severe and critical disease was grouped, for each assay there was a significant association between this group and testing antibody positive: Elecsys^®^ Anti-SARS-CoV-2 (χ^2^=19.03, *p*=0.001), Architect SARS-CoV-2 IgG (χ^2^=15.72, *p*=0.003), Euroimmun SARS-CoV-2 IgA (χ^2^=21.11, *p*=0.007), Euroimmun SARS-CoV-2 IgG (χ^2^=18.77, *p*=0.016), and Trillium IgM (χ^2^=11.59, *p*=0.021) and IgG (χ^2^=17.20, *p*=0.002). Detection of antibodies was highly congruent between assays (Fig. 2).

**Fig. 1.**
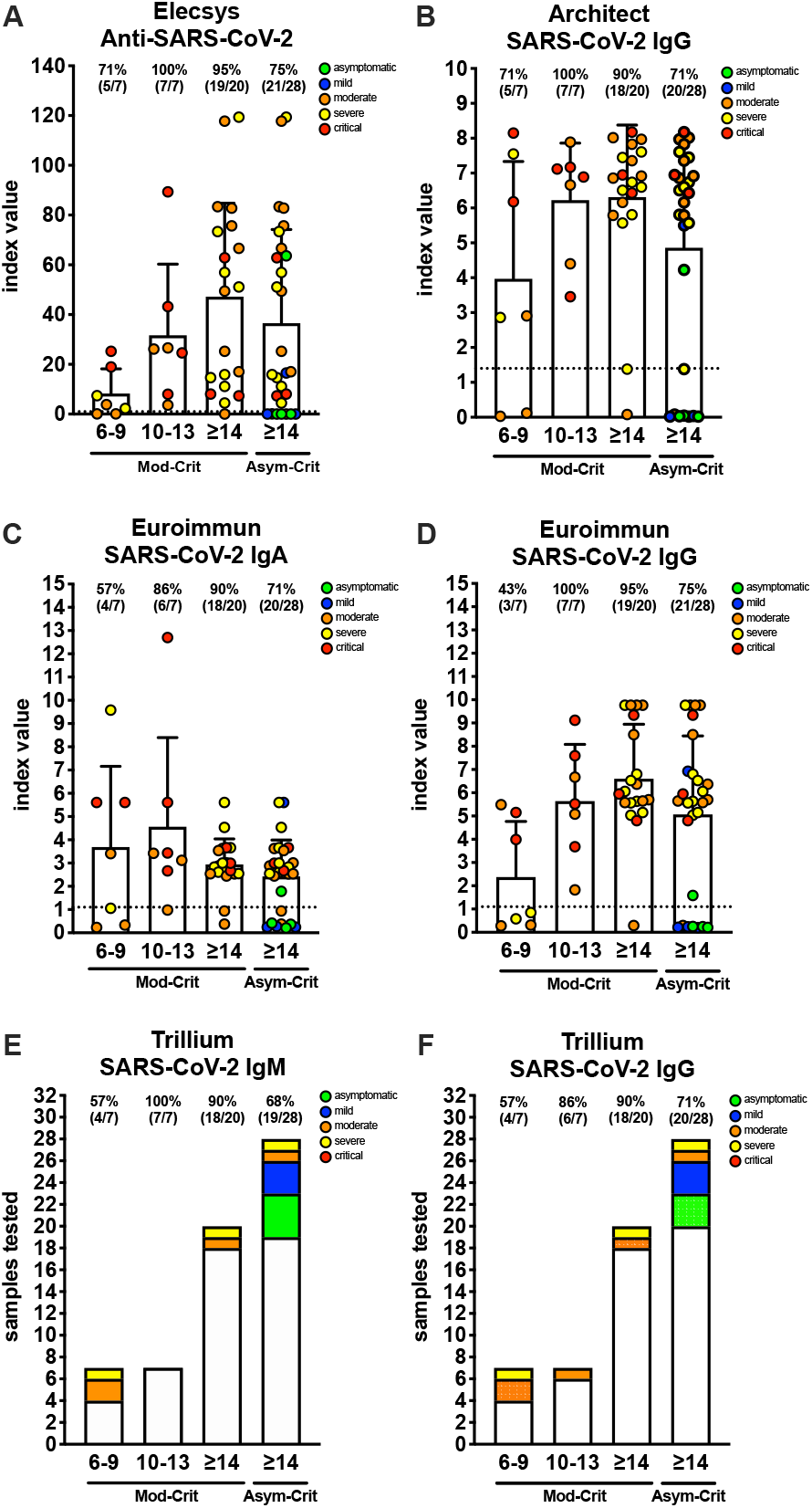
SARS-CoV-2 antibody index values by days after symptom onset for SARS-CoV-2 PCR positive persons. Means with standard deviations are displayed for A) Elecsys^®^ Anti-SARS-CoV-2, B) Architect SARS-CoV-2 IgG, C) Euroimmun SARS-CoV-2 IgA, and D) Euroimmun SARS-CoV-2 IgG assays. Horizontal dotted lines indicate cutoff values. For E) Trillium SARS-CoV-2 IgM and F) Trillium SARS-CoV-2 IgG, white bars indicate the number of positive samples and colored bars indicate samples testing negative. Disease severity is color coded as follows: green = asymptomatic, blue = mild, orange = moderate, yellow = severe, and red = critical.

**Fig. 2.**
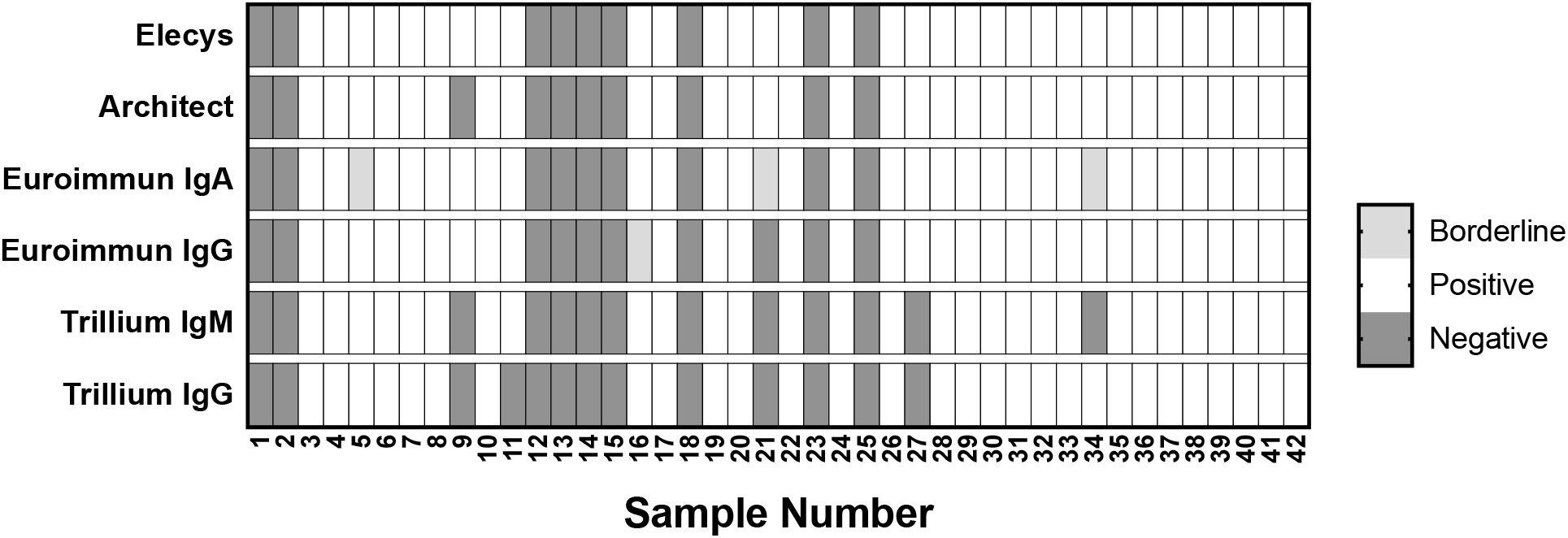
Agreement between SARS-CoV-2 antibody assays. Results for all samples tested for each antibody testing platform are shown. Positive results are shown in white, borderline results in light grey, and negative results in dark grey.

The low diagnostic sensitivity across testing platforms for asymptomatic and mild SARS-CoV-2 infections led us to question whether a relationship exists between the presence of antibodies and relative SARS-CoV-2 viral load at laboratory diagnosis. SARS-CoV-2 real-time PCR cycle threshold (Ct) values were compared with the presence of antibodies from patients with samples collected ≥14 days after onset of symptoms or an initial SARS-CoV-2 PCR. Ct values were significantly different between samples testing positive and negative for all assays examined (Fig. 3). For the Elecsys^®^ Anti-SARS-CoV-2, Architect SARS-CoV-2 IgG, Euroimmun SARS-CoV-2 IgG, and Trillium IgG assays, the mean Ct value was 23.5 ± 5.7 for samples testing antibody positive and 34.6 ± 1.0 for samples testing antibody negative. Mean Ct values for Euroimmun IgA were 24.0 ± 5.7 for samples testing antibody positive 31.8 ± 6.8 for samples testing antibody negative, and for Trillium IgM, 23.0 ± 5.8 for samples testing antibody positive and 33.5 ± 2.8 for samples testing antibody negative.

**Fig. 3.**
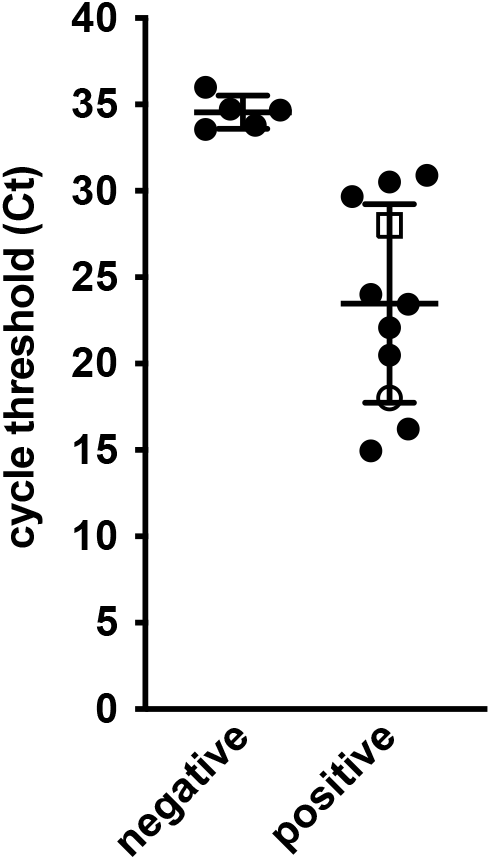
SARS-CoV-2 PCR cycle threshold values for samples testing SARS-CoV-2 antibody negative or positive samples. Sera from SARS-CoV-2 RT-PCR confirmed persons was collected≥14 days after onset of symptoms or ≥14 after a positive SARS-CoV-2 RT-PCR test for asymptomatics. Results were identical for all platforms with the exception of Euroimmun SARS-CoV-2 IgA that tested borderline (open circle) and Trillium IgM that tested negative (open square). Differences between groups was highly significant for all testing platforms. *p*=<0.0001 for Elecsys^®^ Anti-SARS-CoV-2, Architect SARS-CoV-2 IgG, Euroimmun SARS-CoV-2 IgG, and Trillium IgG. *p*=<0.0003 for Euroimmun SARS-CoV-2 IgA and Trillium IgM.

## 4. Discussion and conclusions

Our data examining two chemiluminescent assays, two ELISA assays and one rapid test show that the diagnostic sensitivity of these assays for SARS-CoV-2 antibodies is comparable. Each of the tests examined in this study are approved by the FDA and are CE marked, except for the new Trillium IgM/IgG rapid diagnostic test. The similar diagnostic sensitivity and specificity of the Trillium IgM/IgG rapid diagnostic test with chemiluminescent and ELISA assays makes this test suitable for resource limited laboratories lacking high cost instruments and those that may run laboratory-based assays infrequently in batches.

An accumulating body of evidence indicates that after a SAR-CoV-2 infection antibodies become detectable approximately one week after disease onset [2]. In agreement with these studies, approximately half of the SARS-CoV-2 infected persons in our study had detectable antibodies 6-9 days after onset of symptoms, with most having antibodies ≥10 days after symptom onset.

The high sensitivities for moderate to critical SARS-CoV-2 infections for each of the assays examined is consistent with previous studies [2]. When asymptomatic and mild groups were included in our analysis, sensitivities decreased for all assays. Previous studies have shown that asymptomatic persons are less likely than symptomatic persons of having detectable SARS-CoV-2 antibodies [3,4].

Comparing SARS-CoV-2 antibody results revealed a striking difference in relative viral loads (Ct values) between persons testing antibody negative or positive, with patients testing antibody negative having significantly lower viral loads than patients testing antibody positive. These data are consistent with a recently published study examining SARS-CoV-2-infected asymptomatic contacts and outpatients showing that SARS-CoV-2 real-time PCR Ct values are inversely related to SARS-CoV-2 IgG index values using the Euroimmun SARS-CoV-2 IgG ELISA assay [4]. High SARS-CoV-2 viral loads could cause a more robust adaptive immune response leading to production of high levels and quality of SARS-CoV-2 antibodies.

Our data assessing a Caribbean population of predominantly African descent highlights the limited diagnostic sensitivity of the assays examined for persons with asymptomatic and mild SARS-CoV-2 infections. This finding has important implications for future seroprevalence studies in which a sizable proportion of the SARS-CoV-2-infected population may have experienced no symptoms or mild disease.

## Data Availability

All data is available in the submitted manuscript.

## Declaration of competing interest

All authors declared no conflict of interest.

## Acknowledgements

As a Global Infectious Diseases Scholar, Tiffany Butterfield received mentored research training in the development of this manuscript. This training was supported in part by the University at Buffalo Clinical and Translational Science Institute award UL1TR001412 and the Global Infectious Diseases Research Training Program award D43TW010919. The content is solely the responsibility of the authors and does not necessarily represent the official views of the Clinical and Translational Science Institute or the National Institutes of Health.

## Notes

### Competing Interest Statement

The authors have declared no competing interest.

### Funding Statement

No external funding was received for this study.

### Author Declarations

This study was approved by the UWI Mona Campus Research Ethics Committee (ECP 244 19/20).

